# Post-psychedelic and meditation-related challenges: a protocol for a mixed-methods longitudinal-observational study

**DOI:** 10.64898/2026.10.26.26366468

**Authors:** Joshua Lipson, Nicholas Canby, Zainab Salako, Deanna Kaplan, Santiago Jose Arconada Alvarez, Holly F. Levin-Aspenson, Willoughby Britton, Adie Rae, Todd Korthuis, Nicholas T. Van Dam, Charles L. Raison, Boadie Dunlop, Jessica L. Maples-Keller, Barbara Rothbaum, Roman Palitsky

## Abstract

**Introduction:** Psychedelics and meditation are increasingly used for therapeutic and wellness purposes. However, research into adverse effects (AEs) has been limited, without adequate investigation of the nature, clinical characteristics, and diversity of persisting challenges following psychedelic use and meditation practice. This study aims to better characterize the clinical and contextual features of these challenges, describe their longitudinal course, and compare psychedelic-related challenges to meditation-related challenges, selected as comparator conditions given previously characterized phenomenological and contextual overlap between the two.

**Methods and Analysis:** This is a mixed-methods, enriched-sample, active-comparator (psychedelics vs. meditation) observational study comprising (1) a cross-sectional retrospective survey (N = 1,600), (2) a prospective 12-month longitudinal design involving six bimonthly surveys (n = 800), (3) 12 weeks of weekly ambulatory assessments combining quantitative symptom ratings with narrative voice report (n = 40), and (4) qualitative interviews (n = 60). Participants are recruited into two parallel arms of 800 each: individuals reporting psychedelic-related challenges and individuals reporting meditation-related challenges, defined as distress or impairment persisting after the acute effects of a psychedelic experience or period of meditation practice have subsided. Data collection includes a tailored item pool aimed at capturing the symptoms and phenomenology of these challenges in detail, developed from qualitative research and collaboratively with stakeholders with lived experience. The World Health Organization Disability Assessment Schedule (WHODAS 2.0) and measures of agent/exposure, set, setting, and social response are administered to characterize biopsychosocial correlates and functional impacts of AEs. Initial analyses will describe the phenomenology, course, and functional impact of psychedelic- and meditation-related challenges, and evaluate the structure and psychometric properties of the novel item pool. The study will produce a publicly available de-identified dataset supporting subsequent analyses, such as correspondence with recognized clinical syndromes, trajectory characterization, and cross-arm comparison of measurement structure, reported in registered reports or preregistrations.

## Introduction

### Overview and orientation

This protocol describes an observational, active-comparator naturalistic study of psychedelic-related and meditation-related adverse effects (AEs). Psychedelics and meditation have both shown evidence of efficacy for addressing multiple difficult-to-treat psychiatric symptoms in clinical populations and for enhancing wellbeing in adults (Griffiths et al., 2016; Goldberg et al., 2022; Hoge et al., 2023; Mitchell et al., 2021; Raison et al., 2023). There is also growing evidence, however, that both may produce adverse impacts among a subset of users and practitioners (Breeksema et al., 2022; Simonsson et al., 2023; Goldberg et al., 2022; Van Dam et al., 2025). The therapeutic effects of both have been attributed to alterations in consciousness and heightened neuroplasticity (Holas & Kamińska, 2023; Kishon & Cycowicz, 2025), and although AEs have been recognized in both domains, their nature, chronicity, and correlates remain poorly understood and subject to under-reporting in both the psychedelic (Cheung et al., 2025; Marinis et al., 2025; Olofsson et al., 2026) and meditation (Wong et al., 2018; Van Dam et al., 2025) literatures.

Characterizing unexpected and unwanted effects in psychedelics and meditation is complicated by the fact that both practices can produce intense, difficult, or disorienting experiences during the acute period, many of which resolve without lasting consequence. This protocol therefore focuses on effects with persistent or lasting impact. Throughout this protocol we use *adverse effect* (AE) to denote distress or functional impairment that persists after the acute effects of a psychedelic experience or a period of meditation practice have subsided: challenging, difficult, or distressing sequelae that impact everyday functioning (Britton et al., 2021; Ward et al., 2026). Defined as such, these effects are neither rare nor trivial. Among individuals with a history of psychedelic use across naturalistic contexts—including recreational, ceremonial, and therapeutic use outside of clinical trials—an estimated 6.4–8.9% report difficulties or functional impairment persisting beyond one day (Simonsson et al., 2023; Olofsson et al., 2026), and 1.3% report difficulties lasting more than a year (Olofsson et al., 2026). Comparable figures are observed among meditators: in population-based samples, impairment lasting longer than one day was reported by 3.8–6.0% of participants (Goldberg et al., 2022; Van Dam et al., 2025), with 1.2% reporting impairment lasting one month or longer (Goldberg et al., 2022). Reported prevalence varies considerably, however, depending upon assessment method and definition of AEs used (Britton et al., 2021). A recent review reports that 25–87% of meditators have experienced AEs, according to less conservative definitions, with 3– 37% experiencing AEs that entail functional impairment—though the proportion of these experiencing lasting harms, rather than only transitory difficulties, remains undetermined (Matko & van Dam, 2026).

The remainder of this introduction develops the rationale for the study’s design: the state of research into psychedelic- and meditation-related difficulties, measurement gaps present in both fields, and the logic of using meditation-related challenges as a comparator condition for post-psychedelic challenges, before stating the study’s research questions. A discussion of the state of research in both of these respective areas—as well as their outstanding needs—follows below.

### Psychedelic-related difficulties: the state of research

Classic psychedelics, including psilocybin, lysergic acid diethylamide (LSD), dimethyltryptamine (DMT), and related serotonergic compounds, produce marked, temporary alterations in perception, cognition, and sense of self, and have been investigated as adjuncts to psychotherapy for depression, post-traumatic stress, and substance use disorders (Griffiths et al., 2016; Mitchell et al., 2021; Goodwin et al., 2022). Although they remain Schedule I controlled substances in the United States, several have received FDA breakthrough-therapy designation, are the subject of active Investigational New Drug applications, and their use in the general population has grown substantially in recent years (RAND, 2026). As interest in therapeutic potential and general public use grow in parallel, so do the stakes for a rigorous characterization of the harms that can follow these experiences.

Evidence of such potential for harm has accumulated alongside the evidence of benefit (Breeksema et al., 2022; Simonsson et al., 2023), but its measurement remains uneven. The assessment of acutely challenging experiences during psychedelic sessions is comparatively well-established (Barrett et al., 2016); the assessment of post-psychedelic difficulties, however, remains underdeveloped in two respects. A recent systematic review found that under 25% of psychedelic clinical trials conducted since 2006 used any systematic AE reporting. Furthermore, adverse events in clinical trials have typically been captured through standardized medical-coding frameworks such as MedDRA (Brown et al., 1999), which were developed for conventional drug trials and are poorly suited to describing phenomena that psychedelics may produce, such as subjective and perceptual disturbances, disruptions to the sense of self, and interpersonal or psychotherapeutic sequelae (Palitsky et al., 2024a), and may therefore miss the effects of greatest clinical relevance. Moreover, much of what is known about enduring difficulties outside of clinical trials is based on retrospective self-reports without repeated longitudinal follow-up, such that inference is limited by recall bias and the absence of documentation regarding the timing and context of onset (Shiffman, Stone, & Hufford, 2008).

### Meditation-related difficulties: the state of research

Meditation has yielded a more developed, albeit still limited, literature on AEs. Meditation here refers to a broad family of contemplative practices that train attention, cognition, affect, and self-related processing. Rather than adopting a tradition-specific definition, we characterize meditation in mechanistic terms following Dahl, Lutz, and Davidson (2015), who distinguish attentional, constructive, and deconstructive families of practice, and Dorjee (2016), who frames contemplative practice in terms of the mind’s metacognitive self-regulatory capacity and associated modes of existential awareness. This mechanistic framing spans secular mindfulness-based programs and their contemplative-tradition antecedents. Lindahl et al. (2017) produced a uniquely detailed and standard-setting systematic mapping of meditation-related difficulties, conducted in the Varieties of Contemplative Experience Study. This mixed-methods investigation identified 59 experiential categories of meditation-related challenge across affective, cognitive, perceptual, somatic, conative, social, and sense-of-self domains.

Historically, most quantitative meditation research has assessed adverse experiences using single items or narrow negative-affect indices rather than instruments designed to capture the full range of reported phenomena (Van Dam et al., 2025). Where harms have been monitored in trials at all, monitoring has been inconsistent and adverse-event reporting correspondingly sparse: A systematic review of mindfulness-based stress reduction and cognitive therapy trials found that only a small proportion of eligible RCTs reported monitoring for adverse events at all, and that reported adverse-event rates in those that did were under 1% (Wong et al., 2018). A growing body of multi-item and phenomenological work has begun to address this gap (Britton et al. 2021; Goldberg et al., 2022; Schlosser, 2019; Van Dam et al., 2025), but the evidence base remains small, and prospective data on the emergence and resolution of these difficulties are scarcer. A parallel prospective cohort study now underway in a general population of beginning meditators (Ward et al., 2026) will estimate the incidence and functional impact of these effects as they arise, a design complementary to and coordinated with the present study.

### Meditation-related challenges as comparator condition for psychedelics

Meditation-related AEs overlap with psychedelic-related AEs in both phenomenological and contextual characteristics, which is what makes meditation the most informative available comparator condition (Grob & Grigsby, 2022; Millière et al., 2018; Palitsky et al., 2024b). What has not been established systematically and empirically is the precise nature of this overlap: where these challenges converge in their features, course, and impact, and where they diverge. The present study’s parallel, enriched-sample design is built to answer these questions.

### A need for improvements in measurement and characterization of AEs

Considered together, these two literatures are characterized by a number of common deficits, described in a recent integrative review (Palitsky et al., 2024b), which proposes an agenda for advancing AE research in both domains. Specifically, AE measurement strategies require improvements in detail and comprehensiveness, grounding in the experiences of affected individuals, orientation to real-world impact, standardization, and representativeness. To help address some of these needs, this project aims to improve the content validity (i.e., comprehensiveness, relation to phenomenology, grounding in stakeholder experiences) and advance the methodological rigor of AE assessment related to meditation and psychedelics use.

This study offers considerable granularity in assessing the range of phenomenology that AEs related to these practices may produce, including dissociation, perceptual changes, somatic disturbances, and existential distress—above and beyond the narrow set of general symptoms such as anxiety or depressed mood. It remains unclear whether meditation-related and post-psychedelic challenges constitute variants of recognized clinical syndromes, on one hand, or phenomena not captured by existing nosology, on the other. The assessment approach described in this protocol embeds established measures of clinical symptoms, including depression, anxiety, dissociation, somatic symptoms, alongside novel, phenomenologically-oriented items. This design enables future benchmarking of psychedelic- and meditation-related challenges against recognized syndromes in pre-registered analyses.

There is also a need for chronological assessment beyond cross-sectional self-report, in AE research related to meditation and psychedelics. Longitudinal research methods make it possible to prospectively assess the course, duration, and persistence of these challenges, as well as longitudinal predictors of improvement and worsening. Addressing this lacuna is a central rationale for the longitudinal component of the present design.

### The present study

This protocol describes a mixed-methods, enriched-sample observational study of psychedelic-related and meditation-related challenges, comprising parallel cross-sectional, longitudinal, qualitative interview, and intensive weekly mixed-method diary components (described in **Materials and Methods)**. Notably, several of the symptom batteries and set-and-setting measures were co-developed with stakeholders with lived experience of these challenges and grounded in existing research characterizing AEs (Sánchez et al., 2021; Boness et al., 2025). The study is designed to yield a uniquely detailed dataset characterizing these challenges and their impacts, and to serve as a foundation for a pre-registered program of subsequent analyses. Three sets of research questions guide the initial analyses planned for this study.

The first of these is: What are the phenomenological features of psychedelic- and meditation-related challenges, and what is the structure of these features? Initial analyses will describe the Comprehensive Assessment of Psychedelic-related Challenges (CAPC; see below for description) and Comprehensive Assessment of Meditation-related Challenges (CAMC; see below for description) batteries and evaluate their structure and psychometric properties using exploratory and confirmatory factor analysis.

The second set of questions is: How are these challenges experienced by those affected, what are their impacts on day-to-day life, how do these impacts unfold over time, and to what extent are they associated with functional impairment? These questions are addressed through qualitative interviews, weekly mixed-method diaries collected via a smartphone-administered ecological momentary assessments using Fabla (Kaplan et al., 2025), and the WHO Disability Assessment Schedule (WHODAS 2.0; Üstün et al., 2010).

The third set of questions is: To what extent do psychedelic-related and meditation-related challenges converge or diverge in their features, course, and impacts? The parallel two-arm design permits direct descriptive comparison across all study components.

Additional questions related to this research program will be addressed in pre-registered analyses and registered reports (see Open Science strategy). Beyond these initial and planned analyses, a central aim of this study is to establish a publicly available anonymized dataset intended to serve as a resource for the field. The breadth of assessment described here supports a variety of research questions concerning meditation- and psychedelic-related AEs, their contexts, and impacts The dataset and associated materials will be made available to researchers (see Open Science Strategy).

## Materials and Methods

### Study design

This is a two-armed (psychedelics and meditation) mixed-methods observational study. It comprises a cross-sectional retrospective online surveys (n = 800 per arm; N = 1,600), which serve as the baseline for longitudinal follow-ups among a subset of participants with current unresolved symptoms from recent-onset AEs (n = 400 per arm; N = 800), consisting of six bi-monthly surveys over 12 months. Qualitative interviews will be conducted with a subsample of participants (n = 30 per arm; N = 60), and an additional subset of n = 40 participants drawn from the longitudinal sample will be invited to complete weekly ambulatory assessments comprising quantitative symptom ratings and a narrative voice diary for 12 weeks. Each subsequent component is nested within the preceding one: longitudinal participants are drawn from the cross-sectional sample, and diary participants from the longitudinal sample. presents the study design and timeline.

An enriched-sample design was chosen to support detailed characterization of these challenges. General-population cohort studies can estimate how frequently such difficulties occur, but at realistic incidence rates they yield too few affected individuals to characterize the structure of those difficulties in depth: a cohort of several hundred beginning meditators produces on the order of dozens of adverse-effect cases (Ward et al., 2026), insufficient for detailed analyses of phenomenology or quantitative approaches that can identify symptom clustering. Solely recruiting individuals who have already experienced psychedelic- or meditation-related challenges concentrates the sample on the phenomena of interest and permits the detailed, multi-method assessment described below, although it forecloses analyses of prevalence of these AEs in the general populations of meditators or psychedelics takers.

All components of the study will be conducted online, with surveys delivered via the Qualtrics survey platform, interviews conducted over the Zoom teleconferencing platform, and weekly assessments collected via the Fabla app (Kaplan et al., 2025).

### Participants

This research will recruit N = 1,600 participants into two equal study arms (n = 800 each), identified by primary precipitating exposure (psychedelics vs. meditation).

Inclusion in either arm requires an experience the participant believes was caused by the psychedelic use or meditation practice, and which persisted after the acute effects of the substance had subsided or beyond a single meditation session, and either (1) caused distress, (2) negatively impacted the participant’s life or functioning, or (3) required additional support or treatment. The definition is common to both arms, differing only in the precipitating event.

Psychedelic-arm AEs must be precipitated by either a “classical” psychedelic (5-HT2A receptor agonist), such as LSD, psilocybin, ayahuasca, DMT, 5-MeO-DMT, or mescaline, or by compounds that have been culturally classed with psychedelics, such as MDMA, ibogaine, or ketamine, in line with prior research on subjective and community use features of psychedelics (Palitsky et al. 2024a). Cannabis, though sometimes grouped with psychedelics, differs from the above compounds in its patterns of use, legal status, and neuropharmacology, and carries recognized independent risks associated with its use (Volkow et al., 2014), and is therefore not included. Eligibility for the meditation arm was not limited to any prespecified set of practices or traditions, although these were assessed in the study measures.

Sample stratification targets were developed to support the recruitment of participants with ongoing AEs into the longitudinal study, such that n = 400 participants (50%) in each arm must have experienced <70% improvement in AE symptoms and have had their AEs onset within the past 12 months. For the psychedelics arm, stratification targets also support adequate representation of different psychedelics, comprising at least 150 participants with AEs related to each: psilocybin, LSD, and MDMA. Because of the existing use of ketamine in legal therapeutic settings, its occasional inclusion among psychedelics, and the corresponding need to assess its AEs, ketamine is included in this study; however, because the number of ketamine users may far exceed that of other psychedelics, the stratified sampling approach caps the number of individuals with ketamine-related AEs at n = 150 total. Because other psychedelics (e.g., mescaline) are expected to be used less prevalently, no minimum sampling quota is set for AEs related to these substances.

Participants who fail validity checks or who appear not to be participating in good faith will be excluded. These procedures are described below.

### Recruitment, screening, and compensation

#### Recruitment

Participants will be recruited through several channels. Direct outreach will be made to clinical and academic centers conducting clinical trials, asking them to share study flyers and information with participants who have completed their research studies. Outreach will also be made to community organizations, advocacy networks, and educational entities, some of which provided formative input into the design of this study as part of community-engaged research. These include peer support and special-interest organizations; organizations that support individuals experiencing recent or enduring psychedelic- and meditation-related AEs; and participants receiving psilocybin services at Oregon licensed psilocybin service centers, recruited with the support of community-engaged organizations informed about this study. Further recruitment will occur through public appearances, lectures, and presentations (e.g., podcasts), as well as news bulletins from psychedelic-focused organizations (Palitsky et al., 2025). In the case that recruitment milestones prove difficult to reach, paid targeted social media campaigns may be used for further recruitment. Participants will be recruited from countries where psychedelic use has been decriminalized or legalized in any jurisdiction, pending country-specific approval by the Emory University IRB and international recruitment guidelines; current recruitment includes the United States, Canada, and Australia. Recruitment is planned to continue for 36 months in order to reach adequate sample size and to account for attrition and rapidly changing access to psychedelics internationally.

### Data collection procedure

#### Screening and participant verification

Potential participants for each arm will complete a preliminary screening instrument specific to that arm, querying age, experience of psychedelic- or meditation-related AEs, and willingness to complete a follow-up live screening call. Those who meet initial eligibility criteria will be invited to a live screening conducted over teleconference with study team researchers, typically 15 minutes in length. In this screening, eligibility will be assessed through a set of open-ended questions aimed at discerning participants engaging in research in good faith from fraudulent participants. These will take the form of a free-flowing conversation aimed at eliciting background information that would demonstrate lack of the kinds of experiences that would qualify a participant for this study. These screenings will also provide opportunities to answer detailed questions about the study, offer transparency, and ensure that participants have a direct point of contact with the study team. During study screenings, if participants have had AEs in the context of both meditation and psychedelics, they will be asked to identify one as primary (e.g., due to temporal precedence or subjective impact) and will be enrolled in the corresponding arm of the study. In addition to live screening, survey data will be examined for indicators of non-genuine responding, including failed attention checks, internally inconsistent or impossible responses, implausible response times, and duplicate or automated submissions (Louis & Thompson, 2025; cf. verification procedures described in Ward et al., 2026). Participants identified through these procedures will be contacted when feasible and excluded from analysis if their responses cannot be verified.

#### Survey administration

The baseline survey is expected to take approximately 90-120 minutes. Participants may complete the baseline survey across more than one session, with responses retained and the survey required to be completed within 30 days of starting. All participants will be sent up to three reminder emails and check-ins for each assessment in the study timeline if they have qualified for the study but have not yet begun to participate, or have begun an assessment but have not yet completed it. Follow-up surveys for longitudinal study waves are expected to take approximately 30-60 minutes, with analogous completion windows and check-in procedures as the baseline survey.

#### Longitudinal and weekly diary enrollment

Checks will be conducted weekly to identify participants meeting criteria for longitudinal follow-up (**Participants**). Those participants will be contacted within one week and invited to participate in the longitudinal portion of the study. Invitations will be sent to complete a follow-up survey two months after completion of the baseline survey, and every two months thereafter for one year. Invitations to the voice diary component will be issued on a rolling basis to any participant in the psychedelics arm eligible for longitudinal assessment until all n = 40 diary participants are enrolled; these invitations will be made within the first week after baseline completion, with diaries beginning within two weeks of baseline completion.

#### Compensation

Eligible participants will be compensated with a $30.00 gift card for the baseline survey. Interviews will be compensated with an additional $40.00 gift card. Participants who enroll in the longitudinal portion will be compensated at a progressively increasing rate for each completed assessment, beginning with a base compensation of $10.00 and increasing in increments of $3.00 for each successive wave (i.e., $10, $13, $16, $19, $22, and $25 respectively). Weekly diary participants will be compensated $5.00 per diary entry, totaling $60.00 for 12 entries, with a $20.00 bonus for completing at least 80% of entries.

### Measures

Table 1 presents the schedule of measures by study component and assessment timepoint.

**Table 1.** Schedule of measures by study component and assessment timepoint.

| Measure / assessment domain | Baseline | Longitudinal (bimonthly) | Weekly diary | Interview | Arm |
|---|---|---|---|---|---|
| Individual differences and demographics | ● | — | — | — | Both |
| Psychedelic use history | ● | — | — | — | Psychedelic |
| Meditation practice history | ● | — | — | — | Meditation |
| Agent, Set, and Setting Questionnaire (80 items) | ● | — | — | — | Both; content diverges |
| Influencing factors at onset (timing, health behaviors, circumstances) | ● | — | — | — | Both; content diverges |
| CAPC / CAMC item pool (270 items: including embedded scales such as GAD-7, IDAS-II scales, CAT-PD scales, and others detailed below) | ● | All waves | — | — | Both (variant by arm) |
| WHODAS-II (36 items) | ● | All waves | — | — | Both |
| Overall functioning slider (1 item) | ● | — | — | — | Both |
| Impact and chronicity (ongoing status, duration, recovery slider) | ● | ● | — | — | Both |
| Consequences and service utilization | ● | ● | — | — | Both |
| Appraisals: interpretive frameworks encountered | ● | Waves at 4, 8, 10 months | — | — | Both |
| Responses: disclosure and social reactions (SRQ-S, 16 items) | ● | Waves at 4, 8, 10 months | — | — | Both |
| Institutional Betrayal Questionnaire (up to 53 items) | ● (conditional) | Waves at 4, 8, 10 months | — | — | Both; administered only where professionals or organizations were involved |
| Remedies and supports | ● | Waves at 2, 6, 12 months | — | — | Both; content diverges |
| Multidimensional Scale of Perceived Social Support (12 items) | — | All waves | — | — | Both |
| Spiritual Bypass Scale (13 items) | ● | — | — | — | Both |
| Assessment of causality likelihood (8 criteria) | ● | — | — | — | Both |
| Free-response items | ● | ● | — | — | Both |
| IDAS-II Dysphoria and Well-Being (18 items) | ● | ● | ● | — | Both |
| Sheehan Disability Scale (3 items) | — | — | ● | — | Both |
| Sleep quality (1 item) | — | — | ● | — | Both |
| Voice-recorded narrative prompts (2 items) | — | — | ● | — | Both |
| Semi-structured interview | — | — | — | ● | Both |

#### Baseline survey

Baseline surveys comprise several published instruments, as well as novel items for the assessment of constructs for which published instruments are insufficient or nonexistent. The latter include batteries focused on set and setting, and on the symptoms and phenomenology of AEs.

Participants’ individual-difference and demographic characteristics will include age, dominant hand, gender, sexual orientation, race and ethnicity, national origin and country of current residence, relationship status, rurality, education, current occupation, socioeconomic status (MacArthur Scale of Subjective Social Status; Adler et al., 2000), military service background, religious affiliation, involvement, and observance, and experience as a psychedelic facilitator, guide, or therapist. Participants will also report personal and family psychiatric history. Early childhood adversity will be assessed via the ACEs questionnaire (Felitti et al., 1998; 10 items, yes/no/prefer not to answer).

##### Use and practice history

Participants in both arms report their lifetime history with the relevant practice; items are parallel across arms except where noted. An open-ended item assesses age of first psychedelic use or first meditation practice. Prior motivations are assessed via a 12-item checklist in each arm (e.g., “For social reasons (e.g., my friends were doing it)” in the psychedelic arm; “To gain insight into yourself or parts of your life/self-knowledge” in the meditation arm).

###### Psychedelic arm

The prior psychedelic experiences is assessed via Likert-type items (never, once, 2–5 times, 6–10 times, 11–20 times, 21–50 times, 51–100 times, over 100 times) for each of: psilocybin, LSD, ayahuasca, DMT (other than ayahuasca), peyote, mescaline (other than peyote), ketamine, MDMA, and ibogaine, with three open-ended “other” options.

###### Meditation arm

Prior meditation experience is assessed by asking participants to indicate, from a list of 25 traditions including “no tradition,” which they have practiced in, and from a list of 18 practice types (e.g., “observing the body (body scan),” “embodied movement practice”, “contemplation of sacred texts”), which they have practiced at any point. Participants are then asked to rank the types practiced from most to least, and report cumulative lifetime hours of practice. An additional item asks whether participants have used wearable devices to assist their practice—whatever practice was specified.

###### Both arms

Participants report the contexts in which they have engaged in the practice, using an 8-option checklist (e.g., “At a social event, party, rave, nightclub, festival, or concert”, for the psychedelic arm; “In a meditation teacher training program”, for the meditation arm; “On a multi-day retreat, for both”) with an open-ended “other” option, and rank their endorsed contexts by frequency. Participants who have attended multi-day retreats report how many they have attended and the duration of the longest; psychedelic-arm participants who took psychedelics as part of a spiritual tradition or retreat describe that tradition in an open-ended item. To account for the existence of meditation and psychedelics co-use, both arms query ongoing and historic meditation practice and psychedelics use.

Responses to these items are used to narrow the options presented in the Agent, Set, and Setting Questionnaire below by means of survey logic, such that only previously-endorsed lifetime history responses are re-administered with reference to the index experience.

##### Agent, Set, and Setting Questionnaire

A novel measure for the assessment of the agent, set, and setting at the time of onset of difficulties is included in the baseline questionnaire for both arms. Questionnaire items were developed based on a review of prior literature (Aday et al., 2021; Aixalà, 2022; Breeksema et al., 2022; Carhart-Harris et al., 2018; Haijen et al., 2018; Johnson et al., 2008; Perkins et al., 2021; Russ et al., 2019; Strassman, 2022). The measure comprises 80 items across sections assessing the agent, the individual’s mindset at the time of the index experience (“set“; 36 items), and the context in which it occurred (“setting“; 50 items in the psychedelic arm, 45 in the meditation arm). The full set of agent, set, and setting items is included in **Tables S1 and S2.**

###### Agent

Items assessing the agent focus on the nature of the exposure associated with the onset of difficulties (the index experience).

In the psychedelic arm, participants indicate which psychedelic(s) they took prior to onset, using the response options presented in their use history; estimate how many times they had previously used that compound; specify the method of consumption; and estimate the dose (“Low dose (enough for you to feel a weak psychedelic effect),” “Moderate dose (enough for you to feel psychedelic effects),” “High dose (enough for you to feel extremely strong psychedelic effects),” or “Not sure”), with the option to specify an exact dose if known. Further items ask how frequently they were taking psychedelics at the time, whether they were concurrently microdosing, and whether the index experience was precipitated by multiple doses.

In the meditation arm, participants indicate which of the 18 practice types they were engaged in at the time of onset and the amount of each (from “Very little” to “Very much”), and which was their primary practice, as well as which of the 25 traditions they were practicing in and which was primary, if any. In addition, participants are queried about the typical daily meditation schedule they maintained at the time of the precipitating event; and, where a meditation app was used, its name. The arms differ in one respect of consequence: in the meditation arm, difficulties need not arise from a single, datable index experience, and items accommodate onset that is gradual or cumulative across a period of practice.

In both arms, participants report use of other substances and prescription medications shortly before or during the index experience, including timeline and dosage.

###### Set

Items assessing mindset at the time of the index experience comprise a 5–6 item questionnaire assessing preparatory behaviors, a 12-item checklist assessing participants’ intentions for either psychedelics or meditation (e.g., to improve physical health, for social reasons, general self-improvement, to gain insight into oneself), and two sets of Likert-type items (1 = strongly disagree, 7 = strongly agree) adapted from Perkins et al. (2021). The first set includes 6 items querying mindset around the time of the experience (e.g., occurrence of a recent major stressor; preoccupation with other issues in life; sense of preparedness; mental and physical health; clarity of intention). The second set includes 17 items querying mindset during the experience, assessing perceived control; acceptance or surrender versus resistance; redirection ability; and intense affective states such as fear, despair, and suffering.

###### Setting

Items assessing setting focus on the overall context of the experience (e.g., therapeutic program or therapy, spiritual or religious ceremony or practice, social event; retreat or group-practice context in the meditation arm), the social context (presence of a guide, leader, teacher, sitter, or other participants, the quality of interaction with them, and that person’s expertise or professional role), the environmental context (e.g., safety, comfort, availability of support), the post-experience context (e.g., presence of integration), and additional supports available shortly before or after the experience (e.g., counseling, yoga, tai chi). Items measuring the quality of relationships with others during the experience were adapted from established measures of therapeutic alliance: the Session Rating Scale (Duncan et al., 2003) and the Working Alliance Inventory (Horvath & Greenberg, 1989).

##### Influencing factors at onset of difficulties

Additional factors surrounding the onset of difficulties, which are not properties of the agent, the participant’s mindset, or the context of the experience, are assessed separately and include the following:

###### Timing

Participants report the recency of the index experience and, in the meditation arm, how recently difficulties began, how long after establishing a regular practice they emerged, whether they emerged within the first 50 hours of practice, and how long the daily schedule reported above had been maintained at the time of onset.

###### Health behaviors

Participants in both arms report any changes in diet, food intake, or sleep patterns in the period leading up to the index experience.

###### Circumstances at onset

Participants report their age and relationship status at the time of onset, whether they had children, and whether they were pregnant, had recently given birth, had recently miscarried or terminated a pregnancy, or were trying to conceive.

##### Assessment of symptoms and phenomenology: the CAPC and CAMC

Symptoms and experiences that are part of the AEs are assessed using a novel item pool administered in two variants: the Comprehensive Assessment of Psychedelic-related Challenges (CAPC) and the Comprehensive Assessment of Meditation-related Challenges (CAMC). The two variants comprise the same items, differing in instructions and wording that specify the psychedelic or meditation context. To enable the development and validation of a measurement model from this pool, the CAPC/CAMC is described here as an assessment battery, containing a mixture of validated scales and individual items.

This item pool comprises 270 Likert-type items (1 = not at all, 5 = extremely) designed to capture 59 previously-identified categories of symptoms and experiences across seven domains derived from qualitative research (Lindahl et al., 2017): cognitive, perceptual, affective, somatic, conative, sense of self, and social. It combines 261 items retained or adapted from existing measures, with 9 novel items developed for this study. Items for all CAPC/CAMC scales were either generated in focus groups and/or interviews with meditators and/or psychedelic users with lived experience of enduring negative effects, or were adapted from preexisting scales. Items were rated for inclusion or modification by meditator/psychedelic stakeholders, by researchers from multiple disciplines (religious studies, psychology, etc.), and by a psychometrician in a modified Delphi process (Fink et al., 1984) to yield the final set of items. The full item pool is detailed in **S3** Table.

###### Existing measures included within the CAPC/CAMC

Established measures included within the CAPC/CAMC. Items drawn from existing instruments are listed in an appendix. With three exceptions noted, these instruments are administered in part rather than in whole, and response options were modified throughout for consistency with the rest of the CAPC/CAMC; published subscale scores are therefore not recoverable for partially administered measures, which contribute items to the pool rather than scores. Where an instrument has been further adapted, the adaptation is noted, since evidence of reliability and validity pertains to scores obtained for a particular purpose and population rather than to instruments as fixed properties.

The Generalized Anxiety Disorder Scale (GAD-7; Spitzer et al., 2006) is administered in full and includes 7 Likert-type items assessing symptoms consistent with generalized anxiety disorder. The Inventory of Depression and Anxiety Symptoms–II (IDAS-II; Watson et al., 2012) assesses psychological symptoms across 18 domains, of which 15 are included, yielding 79 items: Appetite Gain (3), Appetite Loss (3), Checking (3), Dysphoria (10), Euphoria (5), Ill Temper (5), Insomnia (6), Lassitude (6), Mania (5), Ordering (5), Social Anxiety (6), Suicidality (6), Traumatic Avoidance (4), Traumatic Intrusions (4), and Well-Being (8). The Panic, Claustrophobia, and Cleaning subscales are not included.

The remaining embedded measures assess loneliness (7 items from the UCLA Loneliness Scale, Version 3; Russell, 1996); dimensions of altered states of consciousness (10 items from the 5D-ASC; Dittrich, 1998); depersonalization (1 item from the Cambridge Depersonalization Scale; Sierra & Berrios, 2000); affective lability, cognitive problems, unusual beliefs, and unusual experiences (28 items comprising four scales of the CAT-PD; Simms et al., 2011); dissociation-type symptoms (8 items from the Curious Experiences Survey, a revision of the Dissociative Experiences Scale; Goldberg, 1999, together with 1 adapted item from the Measure of Dissociative Experiences; Ellickson-Larew, 2019); fear of losing one’s mind (1 item from the Challenging Experience Questionnaire; Barrett et al., 2016); perceptual disturbances persisting after psychedelic use (12 items from the HPPD questionnaire; Locke, unpublished, developed with the Johns Hopkins Center for Psychedelic and Consciousness Research and used with permission); non-ordinary experiences (10 items from the modular Inventory of Non-Ordinary Experiences; Taves et al., 2023); emotional constriction (the 5-item Emotional Constriction subscale of the Multiscale Dissociation Inventory, administered in full; Briere, 2002); mystical-type experience (2 items from the Mystical Experience Questionnaire; Barrett et al., 2015); death-related distress (1 item from the Psychedelic Assisted Therapy Adverse Events protocol; Palitsky et al., 2024a); disturbances in the sense of self and self-boundaries (21 items from the Self and Self Boundary Loss Characteristics Scale; Canby et al., 2024); somatic symptoms (PHQ-15, administered in full; Kroenke et al., 2002); psychotic-spectrum symptoms (4 items from the Psychotomimetic States Inventory; Mason et al., 2008); kundalini-type experiences (6 items from the Kundalini Scale; Thalbourne & Fox, 1999); and unusual experiences (14 items from the item pool developed for the Inventory of Meditation Experiences; Van Dam et al., 2024). Two further items were written for this study adapting the Metacognitive Processes of Decentering Scale (Hanley et al., 2020), and two adapting the Symptom Checklist-90-Revised (Derogatis, 1994).

Finally, the Religious and Spiritual Struggles Scale (Exline et al., 2014) is administered as part of the item pool in a modified 25-item form, with modifications made for relevance to spiritual-but-not-religious participants and to non-Abrahamic traditions (e.g., adding “or a higher power” to items referring to God).

###### Novel items included in the CAPC/CAMC

A total of 9 novel items were developed for this study, derived from input from researchers and stakeholders with lived experience of AEs, and from the 59 categories identified by Lindahl et al. (2017). Example items include “I developed a fear of meditation/psychedelic or meditation/psychedelic-related things” and “I experienced insights about the nature of reality that made me distressed or afraid.“

##### Post-difficulty responses and remedies

Influencing factors occurring after or in response to the onset of difficulties are assessed as follows.

###### Appraisals: interpretive frameworks and explanations encountered

Participants provide an open-ended narrative of their understanding of what caused the difficulties, followed by a checklist of explanatory frameworks they may have encountered from others: scientific or neuroscientific explanations; medical or psychiatric diagnostic explanations; spirituality and religion; trauma-related explanations; spiritual emergency (e.g., kundalini awakening); a normal part of the path or of spiritual progress; spiritual insight; purification (blockages, negative energy, sin, karma, evil spirits); incorrect practice or inappropriate psychedelic use; and pre-existing psychiatric condition, plus two write-in options. For each, participants indicate whether they encountered it and, if so, rate how often (1 = sometimes to 3 = very often) and how helpful or harmful it was (−3 = extremely harmful to +3 = extremely helpful), with optional comments including who provided the explanation.

###### Responses

Participants report whether they disclosed their difficulties to others (e.g., partner, family, friends, community, healthcare professionals, teachers or guides, organizational leadership, “I didn’t tell anyone,” other), with optional comment, and rate the helpfulness of anyone they disclosed to (−3 to +3). A 16-item “Social Responses” inventory, based on the Social Reactions Questionnaire–Shortened (SRQ-S; Ullman et al., 2017), assesses how often participants experienced specific reactions from other people (0 = never, 4 = always). Examples include blame and judgment (e.g., being told they were irresponsible); reassurance and comfort; unwanted control; avoidance; infantilization; and informational support. If participants indicate that helping professionals or organizations in the psychedelic or meditation community played a role, they are administered an adapted version of the Institutional Betrayal Questionnaire (Smith & Freyd, 2013), comprising up to 53 items, which include items centered on psychedelic- and meditation-focused responses that were developed through focus group interviews. These items assess how often institutions or professionals responded in supportive or harmful ways (0 = never, 4 = always). Example responses include active support, treatment as an important member of the community, being warned about risks, being blamed or shamed, being encouraged to view difficulties in spiritual terms, and being discouraged from taking medication.

###### Remedies

Participants report in an open-ended item what they found most unhelpful or harmful, then complete a checklist of attempted remedies: medications; vitamins and herbal supplements; non-prescribed psychoactive substances; psychotherapy; inpatient or residential care; complementary and alternative medicine; meditation or movement-based contemplative practices; psychedelic integration services; lifestyle changes (e.g., diet, exercise, sleep, changes in responsibilities); spiritually oriented resources or organizations; and named organizations (e.g., Cheetah House in the meditation arm). For each endorsed remedy, participants rate how helpful they found it (−3 to +3). Some remedy types elicit further questions: participants endorsing pharmacological treatment are asked which medications they tried; those endorsing complementary and alternative medicine are asked about specific modalities (e.g., acupuncture, energy work, traditional Chinese medicine, Ayurveda, naturopathy, homeopathy, chiropractic). Psychedelic-arm participants are asked about psychedelic-specific coping actions (e.g., changing dose or compound, stopping use, reducing or increasing use, or repeating the same psychedelic experience unmodified or modified); meditation-arm participants are asked about contemplative coping strategies (e.g., attention regulation, acceptance or surrender, self-inquiry, reappraisal or application of views, stopping practice, reducing or increasing practice). In addition, cross-arm use of remedies is assessed: psychedelic-arm participants are asked about contemplative coping strategies for post-psychedelic challenges, and meditation-arm participants are asked about psychedelic use for meditation-related challenges.

##### Impact and chronicity

Participants report whether their difficulties are currently ongoing, how long their difficulties or symptoms lasted (ordinal, 0 = ≤1 day to 8 = >10 years), how recovered they currently feel (slider, 0 = not at all recovered to 100 = totally recovered), and how long they felt impaired as a result (0 = ≤1 day to 8 = >10 years).

###### Consequences and service utilization

Participants endorse any consequences experienced as a result of their difficulties from a list including unemployment or inability to work; leaving school; moving back in with parents; applying for disability or unemployment benefits; suicidal ideation or attempt; urgent care or emergency department visit; psychotropic medication; law enforcement or legal involvement; outpatient care; inpatient medical treatment or hospitalization; and other. Where applicable, number of hospitalizations and total days hospitalized are queried.

###### Functional impairment

An adapted version of the WHODAS-II (Üstün et al., 2010), comprising 36 Likert-type items (0 = none, 4 = extreme), assesses impairment of daily living and functioning over the past 30 days across the domains of cognition, mobility, self-care, getting along with others, life activities, and social participation. Overall functioning is additionally assessed with a single-item slider referring to the period when difficulties were most intense (0 = 0% functioning, 100 = 100% functioning).

##### Spiritual and religious factors

In addition to the measures of religiosity described above, and to the Religious and Spiritual Struggles Scale administered within the CAPC/CAMC, participants complete a modified version of the Spiritual Bypass Scale (Fox et al., 2017), comprising 13 Likert-type items (1 = strongly disagree, 4 = strongly agree). Modifications were made for relevance to spiritual-but-not-religious participants and to non-Abrahamic traditions, analogous to those made to the Religious and Spiritual Struggles Scale.

##### Assessment of causality likelihood

Although self-report assessment of causality is imperfect, this study employs a series of questions across eight criteria designed to address some limitations of self-reported attribution and to evaluate the likelihood that reported experiences were causally related to psychedelics or meditation rather than to alternative causes (Britton et al., 2021; Lindahl et al., 2017). Items query whether pre-existing conditions worsened after exposure; whether experiences occurred on more than one occasion; whether they lessened after stopping or decreasing exposure; whether they reappeared with resumption or increased exposure; whether the participant believes the experiences were caused in whole or in part by exposure; whether experiences first appeared during or following exposure; and whether plausible alternative causes could wholly account for the experience, with an optional open-ended description. This variable additionally serves as a validity control: participants scoring below the cutoff for attribution of their AEs to psychedelics or meditation are excluded from analysis.

##### Qualitative and free-response items

Open-response items appear at regular intervals throughout the sections described above, giving participants an opportunity to comment on any standardized assessment they have just completed. At the end of the survey, participants are invited to submit their full account in writing, along with their understanding of why their experience occurred and which responses and remedies they found most and least helpful. These responses support descriptive characterization and validity checking and may be subjected to content-analytic coding as part of a mixed-methods analytic strategy.

#### Longitudinal follow-up surveys

The longitudinal follow-up surveys, administered every two months (a total of six times) after completion of the baseline survey to participants endorsing ongoing difficulties, employ a subset of the baseline measures: the CAPC/CAMC and the WHODAS-II. They additionally include the Multidimensional Scale of Perceived Social Support (Zimet et al., 1988), comprising 12 Likert-type items (1 = very strongly disagree, 7 = very strongly agree) assessing perceived support from significant others, family, and friends.

At alternating timepoints, participants complete different baseline sections: at 2, 6, and 12 months they complete the Remedies section, tailored to assess what remedies they have currently tried; at 4, 8, and 10 months they complete the Responses and Appraisals sections, tailored to track social responses received during the preceding period.

#### Weekly mixed-method symptom and narrative diaries

A subset of n = 40 participants is invited to complete a weekly assessment for 12 weeks. Diary participants are purposely sampled from the longitudinal cohort on the basis of recency and persistence of difficulties: eligible participants report onset of their index experience within the previous six months and ongoing symptoms that are less than 70% resolved. The assessment is administered via Fabla, a multimedia ecological momentary assessment platform (Arconada Alvarez et al., 2026, Kaplan et al., 2025). Participants receive a brief orientation and app training, including a practice survey and an opportunity to ask questions about the app or the items. Weekly assessments are administered at 7-day intervals at a day and time of each participant’s choosing (**Fig 1**). Each weekly survey remains open for 24 hours, with reminder push notifications delivered at 6-hour intervals until survey completion via Fabla. Adherence is monitored by the research team, with two consecutive missed entries triggering a check-in call.

Each weekly survey takes approximately 10–15 minutes. The first two items are voice-recorded prompts: “Thinking specifically about your [psychedelic *or* meditation] related challenges: how have things been going this week?” and “Tell us, more generally, about your week.” Consistent with prior work using naturalistic narrative speech prompts (Kaplan et al., 2025), participants are instructed to “try to talk about this in a stream-of-conscious way for about 5 minutes. Do not worry about pauses, stutters, or having the right things to say. We are interested in hearing anything about this that comes to mind.” For both narrative diary items, an option to “type my response instead” is provided for participants who prefer to write rather than speak about their experience. Participants then complete the IDAS-II Dysphoria and Well-Being scales (18 items), three items from the Sheehan Disability Scale (Sheehan, 1983) assessing functional impairment, with the total-work-days-lost item removed, and a single item assessing overall sleep quality (1 = terrible, 10 = excellent). In many cases, the stems of these items were modified to query, “During the last week”, given the specific time horizon of this assessment.

#### Qualitative interviews

Participants (n = 30 per arm; N = 60) representing a range of settings, symptom types, use contexts, and compounds, and including participants with both resolved and unresolved symptoms, are invited to participate in qualitative interviews. Interviews use a semi-structured format to evaluate (1) phenomenology, (2) interpretive frameworks, (3) impacts, and (4) contributing factors, including coping strategies, elaborating on methods used by Lindahl et al. (2017) to study meditation-related AEs. Interviews last approximately 60–90 minutes and are conducted after participants have completed the other study components for which they are eligible.

## Analysis Plan

### Scope of analyses reported in this protocol

This protocol describes the study design, the core outcomes, and a defined set of initial descriptive and psychometric analyses, all of which will be preregistered as described in **Open Science strategy**. Beyond data validation and preliminary descriptive characterization of the data, subsequent analyses will be independently pre-registered whenever possible.

### Planned initial analyses

The initial analyses comprise descriptive characterization of the sample and their difficulties, psychometric evaluation of the CAPC/CAMC batteries, description of functional impairment, thematic analysis of the qualitative and narrative data, and side-by-side description across arms. These are the analyses the study will report first; they illustrate what the dataset supports rather than exhausting its analytic potential.

Analyses are to be conducted separately within each arm before any combined analysis. Because psychedelic- and meditation-related AEs are expected to overlap, the effective sample available for evaluating a given item pool may exceed the n = 800 recruited into the corresponding arm; the extent of this overlap is not yet known, and will be reported descriptively.

#### Descriptive characterization

Descriptive statistics will be reported for all study variables: sample composition; the compounds, practices, and contexts associated with onset; endorsement frequencies and severity distributions across the 59 categories comprising the batteries; duration, chronicity, and current status of difficulties; consequences and service utilization; and the remedies participants attempted with their rated helpfulness. Remedy data are characterized descriptively, including whether the profile of remedies attempted differs by the characteristics, severity, or chronicity of difficulties; analyses treating remedies as predictors of outcome are deferred.

#### Structure of the CAPC/CAMC item pool

Analyses specified at this stage of investigation will examine the structure of the CAPC/CAMC item pool. Endorsement rates, variability, and inter-variable associations will be examined first. Then, variables and standalone items showing minimal variability will be flagged and documented, rather than removed. Exploratory factor analysis will then be conducted within each arm, with parallel analysis used to bound the number of factors retained alongside considerations of factor interpretability, followed by confirmatory factor analysis in a holdout sample. Specification of rotation, estimation, and retention criteria will be documented in the pre-registration for this analysis.

#### Functional impairment

WHODAS-II scores will be described by domain and in aggregate, within and across arms, together with the single-item overall functioning rating. Associations between impairment and the characteristics of participants’ difficulties will be reported descriptively.

#### Qualitative analyses

Interview transcripts will be analyzed thematically and using an adaptive, inductive-deductive approach (Layder, 1998). Diary narrative data are automatically transcribed using AWS transcription integration. Transcripts will be checked and corrected as needed by a trained research assistant to verify accuracy, and will be analyzed using an adaptation of the structured tabular approach to thematic analysis for brief texts (Robinson, 2022), suited to the volume and brevity of weekly entries. Narratives and detailed free-responses provided at various points throughout the cross-sectional and longitudinal survey forms may also be retained for thematic analyses.

#### Cross-arm description

Symptom profiles, impairment, course, and remedy patterns will be described side by side across arms, providing the first empirical account of where psychedelic- and meditation-related difficulties converge and diverge. Formal evaluation of whether measurement structures are parallel across arms, in the form of multigroup confirmatory factor analysis and tests of measurement invariance, is deferred, as it depends on the measurement model being established first.

### Sample size calculation

Sample size was determined by the requirements of the full analytic program described in the Open Science strategy below, including analyses to be reported in subsequent registered reports, rather than by the initial analyses alone.

A sample of n = 800 per arm was selected to support the factor-analytic work described above. Current standards suggest validation samples of at least 300 participants for each stage of exploratory and confirmatory analysis (Clark & Watson, 2019); n = 800 permits splitting each arm into training (n = 400) and testing (n = 400) sets while retaining adequate power in both after attrition. Power analysis for regression models indicates that n = 480 provides greater than 95% power to detect small effects (f² of .2). Additionally, this sample size ensures adequate power to support possible future measure-development work, currently beyond the scope of this protocol.

For the longitudinal component, power was estimated through Monte Carlo simulation (Muthén & Muthén, 2002) using the R package *simr*, generating 500 samples of n = 240 to account for 40% attrition, with four repeated measures to account for 33% attrition. Models were fitted using *lme4* and included time, a longitudinal predictor, their interaction, a level-2 variable with a cross-level interaction, and three additional level-2 covariates, assuming random intercepts. Simulations yielded greater than 95% power to detect an effect of .2.

The enriched longitudinal target of n = 400 per arm, with approximately 100 participants at each of 3, 6, 9, and 12 months since onset, was set to provide adequate representation across the range of compounds, practices, contexts, and symptom presentations expected among participants with recent and ongoing difficulties.

### Open Science strategy

The study is registered on the Open Science Framework (https://osf.io/jkxuh/overview), which is the repository of record for the protocol, measures, pre-registrations, analysis code, and amendments. Amendments will be approved by the Emory University IRB, logged on the OSF page, and noted in any publication bearing on affected data. Analyses beyond those detailed above will be specified in pre-registrations or registered reports, which will be solicited for this dataset in advance of data access.

To support secondary use of the dataset by other investigators, study outputs will be made available under a tiered model reflecting the potential re-identification risks of the data types collected. Study materials, such as the protocol, assessment batteries, measure documentation, codebooks, and analysis code, will be deposited openly in the study’s Open Science Framework repository, with analysis code posted at the time of each associated publication. The de-identified quantitative dataset will be deposited following publication of the primary measurement papers, and made available to researchers under a data use agreement specifying non-identification, secure storage, and restrictions on redistribution.

Qualitative interview transcripts, diary narratives, and audio recordings are not deposited: participants disclose use of substances that remain illegal in several jurisdictions, and narrative and voice data carry re-identification risk that de-identification cannot fully eliminate, particularly given the small size of these subsamples. Requests for access to qualitative material will be considered individually, subject to institutional review board approval, an executed data use agreement, and confirmation that the proposed use falls within the scope of participant consent. Requests may be directed to the corresponding author.

## Discussion

### Significance

AEs can occur after both psychedelics and meditation, but their nature is not well understood clinically, hampering efforts to measure, prevent, and treat negative impacts. Current detection paradigms underspecify subjective, SERT, perceptual, self-related, and interpersonal attributes, despite their reported importance to individuals with lived experience of AEs (Argyri et al., 2025; Palitsky et al., 2024a, 2024b). Thus, data that can support an empirically grounded, clinically useful understanding of AEs are needed. This study addresses that need by generating a large, multi-method, longitudinal dataset characterizing these psychedelic- and meditation-related AEs in detail. Complementing qualitative work that has documented AEs falling outside standard clinical-trial detection paradigms (Lindahl et al., 2017; Argyri et al., 2025), it is designed to support the identification of clinically meaningful presentations of psychedelic- and meditation-related difficulties, examination of contributing factors, and detection of factors associated with improvement, persistence, or deterioration, analyses that will be reported in the pre-registered series described above. It further enables the examination of symptom co-occurrence patterns, which can allow for the detection of phenotypes related to psychedelic and meditation AEs, a goal that current AE dictionaries such as the CTC-AE_5 and MedDRA do not readily accommodate.

Difficulties following psychedelics and meditation are sometimes described as elements of an ultimately favorable trajectory, in which challenging experiences carry latent transformative benefits and ultimately yield improvement for those who experience them (Statharakos et al., 2025). This study will generate evidence bearing on trajectories that contribute to benefit as well as to harm, which can allow for an empirical test of the “it gets worse before it gets better” adage. However, rather than categorizing difficulties into harmful or beneficial, the primary focus of this research is to bolster the development of frameworks for supporting individuals who experience AEs, providing care seekers and clinicians with evidence to reduce harm and to support recovery and growth.

The comparison of psychedelic- and meditation-related difficulties across the two arms may help clarify how deliberate practices and psychoactive compounds might differentially relate to subsets of AEs in what is otherwise an often-overlapping corpus (Palitsky et al., 2024b). Nevertheless, the observational design precludes attributing differences to pharmacological action specifically. A clearer account of convergence and divergence will support better detection and response in both contexts.

### Implementation challenges

Several operational considerations bear on execution. The principal challenge is recruiting and enriching the sample to meet the 50% target for recent and ongoing difficulties. Although the study uses an enriched convenience sample, the varied legal status of psychedelics and the often-private nature of meditation practice may produce uneven recruitment rates between arms. Identifying participants whose difficulties began within the past twelve months requires rigorous screening, as many individuals do not categorize their experiences as adverse until considerable time has passed.

The reliance on multiple digital platforms introduces specific considerations for data quality and retention. Given the advertised compensation and an online survey ecosystem saturated with automated and disingenuous responding, the study implements live video screening calls with all prospective participants. Participant burden is also substantial: the baseline survey is estimated at approximately 90-120 minutes. Participants are invited to take breaks and resume at their convenience, and those who do not complete are contacted up to three times to identify barriers and provide reminders.

Longitudinal attrition presents a related risk. With a twelve-month duration and six bimonthly follow-ups, the study is powered to withstand 40% attrition. The team must nonetheless manage the possibility that participants with the highest levels of functional impairment may be the most likely to discontinue, which would bias longitudinal findings; this is partially addressed through personalized contact to identify barriers to completion. Fabla’s design has been purposely built to maintain engagement for long duration studies. A virtual avatar, Fabio, acts as a companion throughout the study nudging and encouraging the participant to complete entries. Preliminary evidence from multiple studies using the app supports that participants really like the avatar element, which paired with additional gamification elements aims to reduce study fatigue. Additionally, the platform enables study coordinators to actively monitor participants and send push notification nudges if entries are being missed. Lastly, a preliminary study comparing Fabla to a text-entry based EMA method found Fabla three times preferable over typed method (Kaplan et al., 2025).

### Limitations

Several limitations attend the design. The first is self-report and retrospective bias: participants may not report accurately on current psychological phenomena (Paulhus & Vazire, 2007), and recollection is influenced by present experience (Ross, 1989). Longitudinal data collection somewhat reduces retrospective and recall bias, though self-report bias remains. The second is the observational design: participants are not randomized, precluding causal inference. The inclusion of a comparator arm helps distinguish effects associated with meditation from those associated with psychedelics, but does not establish causation.

Generalizability is limited linguistically and culturally. Restriction to English-speaking participants in three countries limits inference to other cultural or global contexts in which these practices are common. There is little evidence that the effects of psychedelics, or related AEs, are uniform across cultures, and such conclusions should not be drawn from these findings. Moreover, the enriched sampling design of this study limits inferences about the prevalence of psychedelic- and meditation-related challenges at a population level. Given this sampling strategy, it must be emphasized that participants cannot be taken as representative of psychedelic users or meditators in general.A final limitation is temporal scope. Although one year exceeds the follow-up period of prior studies of psychedelic- or meditation-related challenges, and this study includes more longitudinal assessments than any to date, a one-year window nonetheless restricts interpretation at longer timescales. Longer follow-up would be required to capture the full trajectory of enduring difficulties.

### Risks to completion

The principal risk is low enrollment or high attrition. This is mitigated through a coordinated and flexible recruitment and retention strategy: all research partners contribute to recruitment, progress is monitored regularly, and compensation or procedures are adapted as needed. Participants are clearly informed of study expectations, encouraged to commit to completion, and supported through reminders and follow-ups led by dedicated study personnel. Recruitment is further strengthened by established networks in the psychedelic and contemplative research and practice communities and by structured compensation with bonuses for longitudinal completion. The design accounts for moderate-to-high attrition, permits extension or targeted external recruitment if required, and places key measures early in the battery so that core data are retained even where completion is partial.

### Ethics and safety

Ethical approval has been granted by the Emory University Institutional Review Board (#2025P011692). All participants provide informed consent prior to enrollment.

Because the study involves reporting the use of substances that are currently illegal in several countries, the requirement for written documentation of consent has been waived, on the grounds that a signed consent document would constitute the principal record linking participants to the study and would therefore represent the primary risk of breach of confidentiality. Identifiable information, including email addresses used for longitudinal follow-up, is stored on encrypted servers at Emory University and linked to survey data only via a study identifier. Speech data collected via Fabla are securely and automatically transmitted through HTTPS requests to Emory University’s Health Insurance Portability and Accountability Act (HIPAA)-eligible Amazon Web Services (AWS) infrastructure. Participants retain the ability to review their recordings via the app at all times, and can autonomously choose to delete past voice recordings from the app as well as from research records.

Recruiting a population characterized by distress or impairment requires careful ethical management. The informed consent process and the initial video screening call are designed to give participants clear information and an opportunity to ask questions before beginning the survey, and to establish a direct point of contact with the study team. Participants may skip any item, pause and resume the survey, or withdraw at any point without penalty.

### Study status

At the time of writing, approximately 200 of the targeted 800 cross-sectional surveys have been completed in the psychedelic arm, and approximately 60 of the targeted 800 in the meditation arm. Enrollment into the longitudinal follow-up is underway in both arms. *N* = 9 interviews have been completed for the interview arm of the study. As of the present writing (October 2026), no results have been generated, and no analyses have been undertaken. Participant recruitment and data collection both began in summer 2025; recruitment is expected to be completed by the end of 2028, data collection is expected to be completed by the end of 2029, and initial results are expected by the end of 2028.

### Dissemination

Results will be published in peer-reviewed journals and shared with participating communities, including the peer-support, advocacy, and educational organizations that contributed formative input to the study’s design. Given the community-engaged origins of several study components, findings will be returned to those communities in accessible formats rather than through academic publication alone.

Beyond the analyses reported by the study team, de-identified quantitative data and study materials will be made available to the broader research community under the tiered access model described above, so that other investigators may build on the dataset in future work on psychedelic- and meditation-related challenges. Access to qualitative and audio data is more restricted, reflecting the re-identification risks these data carry.

Registered reports and pre-registrations arising from the study will be posted to the OSF repository as described above.

## Data Availability

Open Science strategy The study is registered on the Open Science Framework (https://osf.io/jkxuh/overview), which is the repository of record for the protocol, measures, pre-registrations, analysis code, and amendments. Amendments will be approved by the Emory University IRB, logged on the OSF page, and noted in any publication bearing on affected data. Analyses beyond those detailed above will be specified in pre-registrations or registered reports, which will be solicited for this dataset in advance of data access. To support secondary use of the dataset by other investigators, study outputs will be made available under a tiered model reflecting the potential re-identification risks of the data types collected. Study materials, such as the protocol, assessment batteries, measure documentation, codebooks, and analysis code, will be deposited openly in the study's Open Science Framework repository, with analysis code posted at the time of each associated publication. The de-identified quantitative dataset will be deposited following publication of the primary measurement papers, and made available to researchers under a data use agreement specifying non-identification, secure storage, and restrictions on redistribution. Qualitative interview transcripts, diary narratives, and audio recordings are not deposited: participants disclose use of substances that remain illegal in several jurisdictions, and narrative and voice data carry re-identification risk that de-identification cannot fully eliminate, particularly given the small size of these subsamples. Requests for access to qualitative material will be considered individually, subject to institutional review board approval, an executed data use agreement, and confirmation that the proposed use falls within the scope of participant consent. Requests may be directed to the corresponding author.

## Supporting information

**S1 Table. Agent, Set, and Setting Questionnaire, psychedelic form: items.** All items administered to participants in the psychedelic arm, with the questionnaire section to which each belongs.

**S2 Table. Agent, Set, and Setting Questionnaire, meditation form: items.** All items administered to participants in the meditation arm, with the questionnaire section to which each belongs.

**S3 Table. CAPC/CAMC item pool and sources.** All 270 items of the Comprehensive Assessment of Psychedelic-related Challenges and Comprehensive Assessment of Meditation-related Challenges, with the source scale and subscale from which each item was retained or adapted, the number of items drawn from each source, and the corresponding citation.

